# “If somebody had told me I’d feel like I do now, I wouldn’t have believed them…” Older adults’ experiences of the BELL trial: a qualitative study

**DOI:** 10.1101/2021.07.15.21260399

**Authors:** Neil J. Meigh, Alexandra R. Davidson, Justin W.L. Keogh, Wayne Hing

**Affiliations:** Bond University, Institute of Health & Sport, Faculty of Health Sciences and Medicine, Gold Coast, Queensland, Australia, 4226; Sports Performance Research Centre New Zealand, AUT University, Auckland, New Zealand; Kasturba Medical College, Manipal Academy of Higher Education Mangalore, Manipal, Karnataka, India

**Keywords:** Older adults, Kettlebell, Group exercise, Physical activity, Healthy aging, Arthritis, Low back pain

## Abstract

**Objectives:** This study examined older adults’ experiences of participating in the BELL trial, involving 12-weeks of group-based hardstyle kettlebell training.

**Methods:** In the BELL trial, 28 insufficiently active older adults (15 women, 13 men, 59-79 years) completed 6 weeks of face-to-face group training, and 6 weeks of home-based training. In-depth semi-structured interviews were audio recorded and transcribed, inductively coded, with themes constructed thematically from patterns of shared meaning.

**Results:** Four higher-order themes were developed that reflect older adults’ experiences participating in a group-exercise program of hardstyle kettlebell training. These included: (1) “It’s one of the best things we’ve done” - enjoying the physical and psychosocial benefits, (2) “It’s improved it tremendously!” - change in a long-term health condition, (3) “It put me on a better course” - overcoming challenges, (4) “I wasn’t just a number” - feeling part of a group/community.

**Discussion:** Findings highlight the perceived physical and psychological benefits of participating in hardstyle group kettlebell training, the value attributed to being part of an age-matched community of like-minded people engaged in group-exercise, as well as the challenges participants faced, and the sense of achievement in overcoming them. Implications for program design and delivery, and future research are discussed.

---

Life expectancy is increasing in many countries worldwide ^(1)^. Recent estimates suggest that approximately 16% of the Australian population are aged 65 or over, with this number projected to increase to 21-23% by 2066 ^(2)^. An aging population and shift in demographic composition, increases the social and economic burden of ill-health, chronic disability, and disease, with the number of healthy years lost to disability, also increasing ^(3)^. Regular physical activity has consistently been found to be protective for older adults facing declines in the domains of physical and cognitive function, self-reported health and vitality, mental health, and mortality ^(4-14)^, with vigorous exercise performed for longer periods providing the greatest benefits ^(15, 16)^. Older adults however, are the least active age group in society ^(17)^, with over 70% of Australian adults aged over 65, insufficiently active ^(2)^. Exercise is the primary means of preventing or reversing the age-related loss of physiological reserve, with frailty accelerated by physical inactivity ^(18)^. Lack of engagement in regular exercise and physical activity is due, in part, to sedentary behaviours being strongly linked with positive reinforcement ^(19)^, thus meaning it is difficult to change public health behaviour. A significant challenge, therefore, is to identify and implement efficacious initiatives that increase long-term physical activity behaviours among older adults. With growing research interest in this area ^(20)^, factors that positively influence attendance rates of physical activity initiatives include; social connectedness, perceived benefits, an enthusiastic and motivating instructor, program design, empowering and energising effects, and being part of a group ^(21-23)^. Engaging with life and self-growth, having a positive attitude, social interaction, and novel pursuits, have been reported by older adults to be valuable components of successful aging ^(24)^. Group exercise in particular is rewarding ^(25)^, and previously trained older adults returning to resistance training describe a renewed enthusiasm for it ^(26)^. Kettlebell training has previously shown to be efficacious for improving function in older adults with Parkinson’s disease ^(27)^, and for improving body composition, strength, and pulmonary function in older women sarcopenia ^(28)^. Data from studies of kettlebell training with younger adults show improvements in dynamic balance ^(29)^, which adds to its potential benefits as a helpful mode of exercise for an older population. Recent findings from the BELL trial clearly support its use for improving grip strength, and measures of health-related physical fitness ^(30)^, but the question remained: would older adults enjoy kettlebell training enough to continue doing it beyond the constraints of a clinical trial?

As a previously untested mode of exercise for otherwise healthy community dwelling older adults, the BELL Trial was conducted to test the effectiveness of group-based hardstyle kettlebell training to promote healthy aging to insufficiently active older adults. Thus, in this study, we explored the experiences of participants in the BELL trial. The purpose of the study was to gain a greater understanding of the participants’ experience of performing hardstyle kettlebell training. The experiences of these participants may provide important insights that can be used to inform future provision of community-based group kettlebell training programs for older adults.

## 1. Methods

The study was approved by the Bond University Human Research Ethics Committee (NM03279), with the design, conduct, and reporting of this study adhering to the Consolidated criteria for reporting qualitative research (COREQ) guidelines ^(31)^. The BELL trial was pre-registered on the Australian New Zealand Clinical Trials Registry (ACTRN12619001177145).

### 1.1. Participants

Thirty-two apparently healthy but insufficiently active older adults (59-79 years) were recruited to complete 12-weeks of moderate to high-intensity hardstyle kettlebell training. Three participants withdrew during the control period: medical condition (*n* = 1), injury (*n* = 1) and no longer able to attend (*n* = 1). Twenty-nine participants commenced training, with 28 completing six weeks or more. Purposive sampling ^(32)^ was used to capture the experience of all participants who completed at least half (≥ 6 weeks) of the training intervention (13 men and 15 women, 68.8 ± 4.6 years). One male, who withdrew in the second week due to work commitments, was not invited for interview.

### 1.2. Study setting

Participants attended 45-minute group-exercise classes three-times weekly (Mon, Wed, Fri), and completed prescribed home exercise twice-weekly (Tue & Thur). Details of the training intervention have been published elsewhere ^(30)^. Face-to-face group classes were conducted by the first author for six weeks, then remotely thereafter due to COVID-19 restrictions. The first author is a 45-year-old male kettlebell instructor and Physiotherapist. Several strategies were used to enhance participant engagement, including frequent individual and group encouragement (recognition of overcoming challenges, extraordinary effort, and achieving a ‘personal best’), daily communication via a private Facebook group and email (to foster a spirit of group support, accountability, camaraderie, and healthy competition), and encouraging post-workout gatherings (on-site café) to promote social connection outside of class. Additionally, the instructor took part in the training where possible, to be seen as an active part of the group experience.

The BELL trial was conducted over a period of 8-9 months. This provided the researcher with a unique opportunity to interact with the participants, and for thoughtful and reflexive engagement in the qualitative data which were subsequently collected. One in three participants who commenced training in February 2020, continued to train together beyond the intervention period (June 2020), and were still training weekly at the time of pre-print publication. The first author remained in regular contact with the group to the time of publication.

### 1.3. Procedures

Data collection took place 6-15 May 2020, following completion of the intervention period 1 May. Participants were invited to take part in an in-depth semi-structured interview, for up to an hour. Interviews were held via Zoom due to COVID-19 restrictions, with the participants at home. Interviews were conducted by the first author who, at the time of the study, was a doctoral student responsible for the BELL trial. Interviews were digitally recorded and transcribed using Trint transcription software ^(33)^. All participants in the study were interviewed separately, and no eligible participants declined to be interviewed. There were no repeat interviews. Given the relationship between the interviewer and participants, interviews were not piloted, and field notes were not taken. Transcriptions were checked against the audio recordings line by line, with errors corrected as necessary before import to NVivo 12 software ^(34)^ for coding and thematic analysis. Transcripts were not returned to participants for comment, however authenticity of quotes was maintained ^(35)^.

An interview guide (see Table 1) was used as a framework however, questions were posed flexibly in an open-ended way, allowing key experiences to be discussed. Participants were given the opportunity to talk about topics which were not part of the guide, allowing space for unexpected insights to be explored ^(36)^. Interviews were informed by the participant’s responses to a series of quantitative rating-scale questionnaires, which had been completed via Survey Monkey following cessation of the training period. Questionnaire responses highlighted areas of interest, allowing the interviewer to ask targeted questions to gain a better insight of meaning, and discuss key topics. Questionnaires were developed based upon the first author’s disciplinary expertise, and experience working with the participants during the training period. Overarching topics were, i) participant’s experience training with kettlebells, ii) perceived positive and beneficial effects, and iii) negative and undesirable effects. *Questionnaires are available as supplementary data*.

**Table 1.**
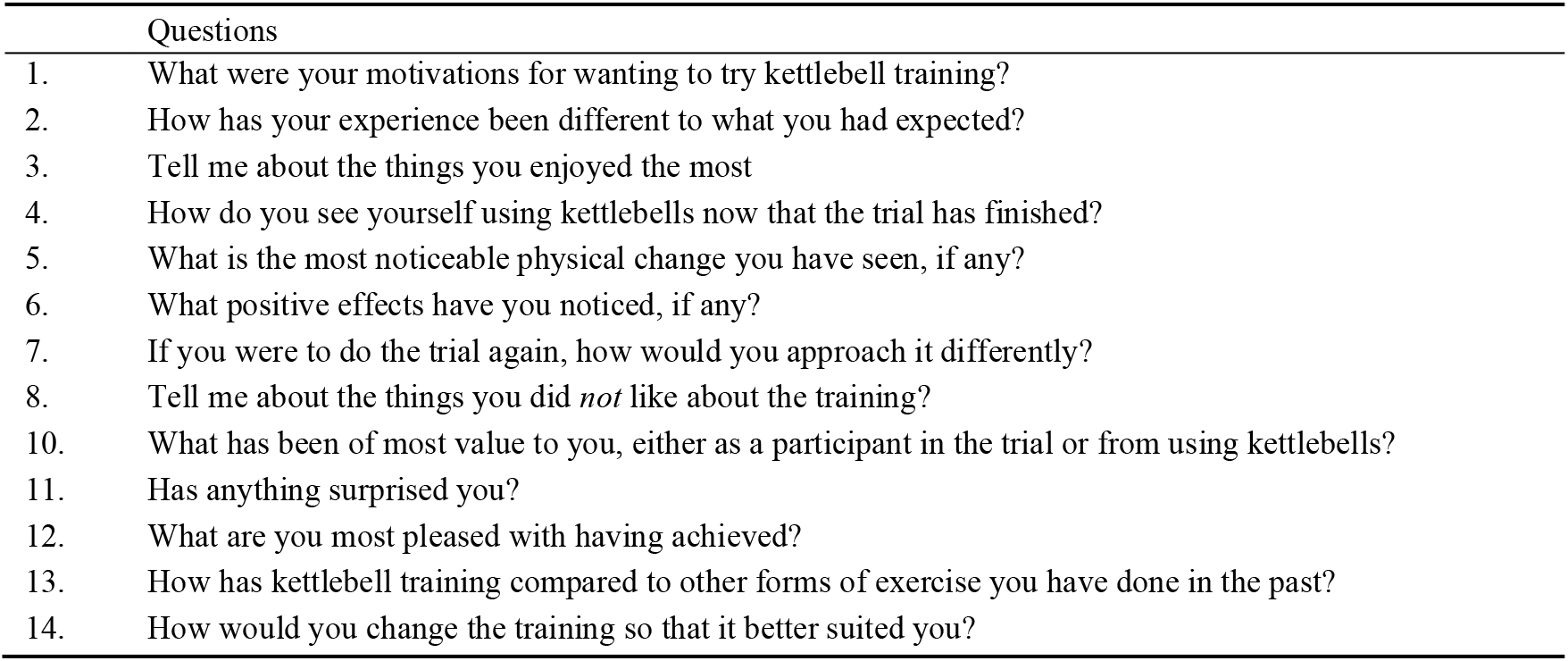
Interview guide

|  | Questions |
| --- | --- |
| 1. | What were your motivations for wanting to try kettlebell training? |
| 2. | How has your experience been different to what you had expected? |
| 3. | Tell me about the things you enjoyed the most |
| 4. | How do you see yourself using kettlebells now that the trial has finished? |
| 5. | What is the most noticeable physical change you have seen, if any? |
| 6. | What positive effects have you noticed, if any? |
| 7. | If you were to do the trial again, how would you approach it differently? |
| 8. | Tell me about the things you did *not* like about the training? |
| 10. | What has been of most value to you, either as a participant in the trial or from using kettlebells? |
| 11. | Has anything surprised you? |
| 12. | What are you most pleased with having achieved? |
| 13. | How has kettlebell training compared to other forms of exercise you have done in the past? |
| 14. | How would you change the training so that it better suited you? |

### 1.4. Philosophical underpinnings and data analysis

This qualitative study, which was subsumed within a larger pragmatic controlled trial, is underpinned by a philosophy of critical realism (CR) ^(37)^, acknowledging the limitations of positivism and constructivism within social science research ^(38)^. To examine older adults’ experience participating in group kettlebell training, data from in-depth, semi-structured interviews, were thematically analysed, with codes and themes developed inductively using reflexive thematic analysis ^(39-42)^.

The data analysis was led by the first author, who had conducted the training intervention with the study participants. Familiar with the lived experience of the participants, the first author began by immersing himself in the data, through reading and re-reading transcriptions of the audio recordings. Consistent with CR ontology, initial data analysis and coding began with a search for ‘demi-regularities’ at the empirical level (the participants description of *events*) and identification of *tendencies* (trends or patterns acting as causal tendencies) ^(43)^. The first round of coding utilised a combination of coding categories, such as emotion (participants’ feelings e.g., *“fun”*), value (values, attitudes and beliefs e.g., *beliefs of effect* or *experiential value*), narrative (participants’ stories e.g., *social interaction*), evaluation (assigning judgement e.g., *increased self-confidence*), and process coding (observable activity e.g., *walking more*), as well as theming data (meaning e.g., *grocery shopping has become easier*) ^(44)^. Codes and initial themes from collated data were developed iteratively, both semantic and latent, until the entire data set was complete, resulting in the construction of 21 broad themes. The focus was to identify patterns of shared meaning within the participants’ accounts of their experience in the trial.

The first and second authors met to review and further analyse the codes developed by the first author, to refine, define, and name higher-order themes. The second author is a doctoral student whose study exclusively utilises qualitative research methods. She had not been involved with any other part of the BELL trial and did not meet the participants. Second round coding methods included pattern (e.g., *identifying trends and relationship*), focused (e.g., *most frequent*), axial (e.g., *identifying core categories and dimensions*) and selective coding (e.g., *connections to form a storyline*) ^(44)^. Transcripts were reviewed to refine the themes, and to choose quotations to support interpretations.

Themes were subsequently reworked and redefined during the writing process. After the main findings had been identified through coding, *abduction* (theoretical redescription) was used to re-describe and raise the theoretical engagement beyond thick description, using general ideas about exercise and resistance training, community-based group programs with older adults, and kettlebell training ^(43)^. Finally, *retroduction* (inference of causal tendencies) was used to describe the participants’ experiences ^(43)^. To maintain a commitment to complexity, and highlight some of the challenges of delivering an exercise intervention to a diverse group of older individuals, the analysis includes exceptions to central themes and discussion of their potential impact ^(45)^. Final higher-order themes are presented in the findings below.

## 2. Results

Four themes were developed that describe how the participants perceived and experienced the BELL trial: (1) *“It’s one of the best things we’ve done”* - enjoying the physical and psychosocial benefits, (2) *“It’s improved it tremendously!”* - change in a long-term health condition, (3) *“It put me on a better course”* - overcoming challenges, (4) *“I wasn’t just a number”* - feeling part of a group/community. Pseudonyms are assigned, and sex has been changed in some cases, to protect participant anonymity.

### 2.1. “It’s one of the best things we’ve done”: Enjoying the physical and psychosocial benefits

Participants described an improved sense of self-confidence from feeling fitter and stronger.

*“I’ve got a pool so I always keep a few 20 kilo bags of salt around the place. I throw that around now like a bottle of milk… our coffee table is really heavy and I just pick one end of it up [to vacuum underneath]… my wife won’t let me tie up the garbage bins with the bags [anymore], because when I tie them up now in a knot I tear the bag completely!”* Samuel, 70-74 years old

Participants reported being more engaged in incidental physical activity, and less breathless with prolonged exercise such as long walks and climbing hills. Participants were happy and excited to be re-engaging in physical activities which they had previously cut back or stopped doing altogether, and being more active with their partners. Domestic activities, particularly those involving relatively heavy objects such as carrying shopping bags, moving furniture to clean, and gardening, were said to have become much easier.

Participants described feeling more energetic and motivated. They enjoyed having goals to aim for, and felt proud when they were able to reach and exceed them. Participants liked the discipline of having to be somewhere and do something every day, and appreciated that it was “*fun*”, with an opportunity to meet people and make new friends. Participants felt that the training had changed their appearance, describing muscles in their arms and legs has having more tone. They described how others in the group appeared to be walking taller, and family members had noticed that they were carrying themselves differently and doing more around the home.

*“I’m walking, going out with the wife, doing things, and I’m confident in myself as I walk around that I’m not an ‘old’ person, I’m [an older person] that looks healthy and I’m quite enjoying that. …Having the opportunity to do something, seeing it through, finishing it, being able to challenge myself; I’m a different [man]”* Alan, 70-74 years old.

Participants said they were happier and enjoyed learning something new. They liked that the training was challenging, the competitiveness of trying to better their own achievements, and comparing themselves to the performance of others in the group. Competition and goal-attainment gave a sense of accomplishment, and participants enjoyed being able to record and track their progress. Participants liked that training sessions were always different and said that not knowing what to expect each class had kept it interesting and made them feel engaged. Participants joked about “*old*” stereotypes and described specific components of the program which had given them a great sense of accomplishment and pride, and reinforced feelings that they were still very physically capable: learning exercises they perceived to be too hard, being able to do things which they thought would be beyond their capacity, getting to the end of a hard training session, and attaining a new ‘personal best’.

Participants said they could see changes in their physical health and had an improved sense of wellbeing. They described being more energetic during the day, sleeping better at night, and liked feeling healthier. Participants said that exploring their physical limits with heavy weights and pushing themselves hard during class, and at home during the final week, had been empowering. It gave them the confidence to know that they could handle day-to-day activities, when previously they may have been anxious about them or avoidant. Several participants said their balance had improved, including one participant whose balance had been adversely affected by chemotherapy, and this had increased their physical confidence during day-to-day tasks, such as walking up and down stairs.

*“Just having a shower, I used to get tired, and as for getting dressed, … I don’t have to hang on to something so I don’t fall over. … cutting my toenails, I never used to be able to do that by myself. …I used to tire out walking up a set of stairs, but now I can do that easily… It’s just been unbelievable the difference in myself. … for me to achieve what I’ve been doing being a smoker, well I just can’t be happier. It’s just unbelievable what I can do. … If somebody had told me I’d feel like I do now, I wouldn’t have believed them, … It’s just made me so much [more] enthused, like if I want to go across the road to get the paper, I wouldn’t ever think about jumping in the car now, I’d either hop on my bike or walk because you can do that now, … it’s one of the best things we’ve done… I feel so much stronger*.*”* Peter, 70-74 years-old

Participants described being more flexible, and how activities such as rising from a low chair had become easier. These improvements in physical function made participants feel good about themselves, which had been reinforced when friends and family noticed the positive changes, or friends their own age told them they couldn’t or wouldn’t be able to do what they were doing.

*“I’ve got a lounge chair at home which is relatively low and I purposely hop up from that chair by not using [my] arms… I put my arms out in front of me and stand up from below normal seat height and I do that every time now and I haven’t got a problem with it. Before, I’d either have to push myself up, or turn over to one side and then stand up. I don’t do that anymore”*. Thomas, 70-74 years old

### 2.2. “It’s improved it tremendously!”: Change in long-term health condition

Many of the participants described how the training had positively influenced a long-term health condition, which had negatively affected their life or physical function. These conditions included hip and knee arthritis, persistent non-specific low back pain, osteoporosis, diabetes, peripheral neuropathy, cancer, ankylosing spondylitis, migraines, alcohol dependency, immunosuppression, obesity, hypertension, hypercholesterolemia, depression, and poor sleep.

All participants who said they had painful arthritic knees described a significant reduction in pain after the trial. In a few cases, participants said their symptoms had initially been irritated or worsened by some exercises, such as lunges, but all the participants who had been living with sore knees reported significant improvements in their arthritic knee pain and function at the end of the trial:

*“Since I adjusted the exercises, I’ve been enjoying it because I don’t have the pain. If you’re enjoying doing it, it’s a motivation to do it… the pain abated, and I still got fit, so it was good”*. Spencer, 65-69 years old

Participants either adapted an exercise so that it was more comfortable, or they chose to push through discomfort regardless. One participant with knee arthritis said he no longer had any pain.

*“I can carry three loads of groceries up the stairs and… they’re quite steep, whereas before with my knee I couldn’t do it”*. Abigail, 65-69 years old

Participants described significant reductions in other painful conditions. One participant, who consistently performed considerably more home exercise than required, had been living with frequent migraines, pain associated with an inflammatory arthropathy, frequently had insufficient sleep, and had a history of trips and falls. After the trial, she reported less frequent headaches, less pain, improved sleep, and better balance:

*“My migraines, they’re less frequent and less severe… to me, wellbeing is not feeling sick with a headache or not having the same nagging pain in my hips… [my] overall well-being is much better… I used to have to go and have massages nearly every week, and I haven’t been once since I’ve been doing kettlebell [training]… I’m not constantly looking down to see that I’m not going to trip over… I’m not feeling as unsteady… I’ve had such a good response to it… I sleep like a baby”* Janice, 60-64 years old.

Several participants had also been living with persistent low back pain. Except for one individual with a 50-year history of back pain who withdrew from the trial, everyone living with back pain described significant improvements in their symptoms and function.

“*When I was getting out of bed before I started the trial, [I] always worried about [my] back going or something, but now I just can’t believe how much stronger my back is now. I’ll get up in the morning, get out of bed and there’s just not one bit of pain in there… It’s definitely one hundred percent better than what it was before I started the trial. It’s just unreal, I can’t believe it*.*”* Robert, 70-74 years old

“*I’m thrilled! I’m fit as a fiddle, I feel good, I sleep well, and I have no more back problems. I used to have once a month a physio come here to maintain my back [ ] I think I don’t need him [any more], I’m fine*.*”* Georgina, 70-74 years old

Participants described feeling mentally buoyed by their participation in the trial. The profound positive psychological impact that engagement in the trial had, was most powerfully described by one participant who had been living with a mental health disorder for 20 years:

*“This was the challenge I need… it’s given me the motivation to continue and that’s sort of saving me… this has been brilliant… I’m actually scared coming out the other side of this… I’m still in a bit of trouble…I have to keep going with this otherwise I’ll go back to where I was last year and not in a good place”*. Katherine, 70-74 years old

### 2.3. “It put me on a better course”: Overcoming challenges

Participants living with pain commonly held maladaptive beliefs and behaviours relating to perceived risk of harm, believing they might hurt themselves if they pushed too hard, lifted too much, or did something which was too uncomfortable. Participants described the training as being harder than they had expected, at times uncomfortable even painful. Swinging the kettlebell overhead to perform a snatch was difficult to learn and created a sense of fear and apprehension among participants who found it difficult or impossible to press an 8kg kettlebell overhead. But the participants persevered with the training, improved their technique, worried less, and even learnt to enjoy the some of the things they had found to be most challenging at the start.

*“When you said we’d be swinging this thing above our head, everybody’s like ‘well I won’t be doing that’ and ‘that won’t be me’, and then when we all did it, it’s like, ‘we can do this, we’ll just keep on going’. That was good; that you got us to do something that I think we were all a little bit hesitant about, [but] by the end of it, that’s my favourite thing, I love it”*. Irene, 60-64 years old

Participants found it was not easy to get up and down from the ground to perform a Turkish get-up, describing insufficient lower limb strength, not having enough flexibility, or having sore knees. Participants who really struggled with the Turkish get-up it, or said the lunging movements irritated their knees, found it demotivating. But they persevered and found ways to make it easier, so that they *could* do it. The Turkish get-up pattern was widely recognised as a floor transfer. Participants saw it as having more value than some of the other exercises because they could see how they were able to use it at home. One participant described how he had jumped onto a kitchen benchtop and performed a Turkish get-up to change a lightbulb. Participants described an increased sense of physical self-confidence having practiced and improved their ability to stand up from the ground.

*“When we first had to do those … I thought I was slow as molasses. I had an ache and pain here and there and now I find myself using the Turkish get-up if I’m sitting on the floor”*. Lucy, 65-69 years old

Delayed onset muscle soreness (DOMS) affected most participants, particularly in the first few weeks, sometimes occurring after exercises described as “*easy*”. Many participants were unfamiliar with the sensation of DOMS and unprepared for how it might impact them. In some cases, participants were reluctant to move much because of the discomfort, sufficiently concerned to describe it as “*severe*”. None of the participants allowed muscle soreness to prevent them from fully participating or allowing it to get in the way of their continued engagement with the group. Participants with a history of using resistance exercise were much less affected by muscle soreness and generally undeterred by it. For some participants, muscle soreness was seen as positive because it made them feel like they had done something which was “*working*”, and it brought back happy memories of being younger and more physically active. When aches and pains occurred, participants remained stoic and carried on regardless, describing a shared commitment to the trial and a determination to reach the end. Participants wanted to overcome their challenges and felt proud achieving what friends and family often told them they shouldn’t be doing, because it was perceived as unsuitable for older people.

*“[I thought to myself] that was too easy, am I doing it correctly? … how hard can it be, you’re just stepping up and down. I just kept going and I couldn’t move the next day. It was really bad [laughing]… It was so easy to do I thought this is nothing. It wasn’t until the next day, I thought holy cow! The [DOMS in my] calves on both legs was shocking, I could hardly move”*. Iris, 65-69 years old

Participants were concerned about hurting themselves, especially if they had a history of back pain, knee arthritis, a heart condition, or maladaptive beliefs about lifting and back injury.

*“The fear [that] people my age have about injury is paramount. We all like to be brave and like to think that we can do things, but sometimes either we don’t do as much as we should and can [do] for fear of hurting ourselves. We just need that extra encouragement to realise, yes, you can do this and it’s okay”*. Kathy, 65-69 years old

Regardless of their concerns, participants continued to push themselves very hard. This was typified by ‘Ken’ who had previously been a competitive sportsman. Ken accumulated, by a very large margin, the highest training load volume in the group. He was so motivated and capable that he felt he was not being pushed hard enough: *“at times, I just felt I was held back a bit with what I wanted to do”*, and later went on to comfortably perform two-handed swings with a 40 kg kettlebell at home. But, after three decades of occupational health and safety training with his employer, Ken started the trial anxious that poor core strength and technique just leaning across the bed to pull up the doona (duvet) might hurt his back:

*“I was really hesitant beforehand. I think I said to you very early, ‘aren’t we going to hurt our backs doing this?’ and you said ‘no’. … That’s been on my mind all the way through; at some stage I’m going to do my back, but I never did. … now there’re no inhibitions with what I’m doing, I’m confident … that I’m not going to hurt myself”*. Ken, 70-74 years old

The training created numerous physical, psychological, and emotional challenges for the participants, to the extent that some thought about quitting in the early stages. However, no participants quit for any of these reasons, instead choosing to persevere and overcome them. A strong sub-theme, which overlapped with the participant’s enjoyment of the program, was the instructor’s role in facilitating this collective can-do mindset. Participants reported that the instructor’s personality, enthusiasm, leadership, encouragement, and happy, helpful demeanour, had been motivating and that was why they had continued to push themselves as hard as they had, often despite obvious challenges.

*“I valued ‘you’ the most, because the time and effort that you put into all of us was awesome, and just the confidence that I had that you knew what you were talking about and that you cared about what we were doing and how we were doing it… I had a lot of confidence; that to me was the most important thing”* Janice, 60-64 years old.

The participant’s attitude toward overcoming challenges was exemplified by Sean, who had experienced persistent shoulder pain for more than half of the trial. During the trial he was also diagnosed as iron deficient, told he had arthritic knees bad enough to warrant surgery, that a rotator cuff problem was the cause of his shoulder pain, and needed surgery to correct cataracts:

*“I was starting to I think I was getting old and decrepit and demented, and that (training) picked me up out of that black hole… you got us all going; it’s your personality… that made me go harder than I would have… I was so buoyed because of the improvement in my physical health… It put me on a better course going into my older years that’s for sure*.*”* Sean, 70-75 years old.

The spirit of the group was perhaps best displayed by Samuel who, in his 70s, had volunteered to participate shortly after a life-changing conversation with his doctor. After his GP had questioned why he wanted to participant in the trial, Samuel told his doctor that he just wanted to be as healthy as he could be. Facing significant adversity, Samuel successfully completed the trial describing remarkable changes in his physical and mental health, all while performing swings at home with a 36kg kettlebell.

### 2.4. “I wasn’t just a number”: Feeling part of a group/community

The group dynamic was one of the strongest perceived benefits for participants. Being among people of a similar age, with varying abilities, provided participants with a rich and rewarding social connection. The camaraderie was fun and enjoyable, and allowed participants to support and encourage one another to succeed and push themselves harder than they would otherwise have done on their own. The group environment, where everyone was starting as a beginner, also gave participants the opportunity to try something new. Participants were more likely to engage in structured exercise if it was with others, and felt part of a positive, supportive, and encouraging community. Andrew, who described himself as a loner, said:

*“In the group, I found that I was more open to being nicer and more inclusive with more people, so it really taught me that I can be part of the group and enjoy people*.*” Andrew, 65-69 years old*

Men described being competitive more frequently than women, and competitiveness was regarded as beneficial, irrespective of how well someone was perceived to be doing in training. For example, those who were doing “better” typically enjoyed being able to help their partners when they were struggling, while those “*chasing*” the leaders, appreciated seeing someone their own age doing better than them, as it gave them something to aim for; *“if they can do it, then so can I!”* Participants especially enjoyed activities performed in pairs, as this allowed those who had formed social friendships outside of the gym to work together. The value of the group dynamic was typified by Sandy who, after a short period of absence due to back pain, was worried that others had progressed without her, and that she may not be accepted back into the group:

*“They were all so happy to see me and it just made me feel that I wasn’t just a number; I was appreciated and that everyone cared how I was, and they didn’t [even] know me very well, … it just made me feel good, … I felt more positive about coming back, … [it] just made me realise how much I was enjoying it, … coming back, … made me realise how good I had been feeling, … I suddenly realised there were so many positive things that I’ve been experiencing that I’d overlooked and then when I got back, I realised, ah, this is what I was actually experiencing*.*” Sandy, 60-65 years old*

The value of the social connection which the group provided, was most profoundly felt when face-to-face training was cancelled due to COVID-19 restrictions. Participants missed seeing the people they had formed new friendships with, and found it difficult to maintain the same level of motivation to train at home. This was somewhat mitigated by a private Facebook page, which allowed people to connect on a social level and share their progress, watch video recordings and live streams of training sessions, and take part in frequent ‘challenges’. The value of the social connection and support was perhaps most poignantly expressed by one participant whose partner was very unwell throughout the trial: *“the contacts and friendships I’ve enjoyed from your trial have been and still are the most beneficial and a comfort to me”*, Jennifer 65-69 years old.

## 3. Discussion

The purpose of this study was to investigate the experiences of insufficiently active older adults, who participated in 3-months of moderate to high-intensity group-based hardstyle kettlebell training. The findings of this study, highlight some of the challenges and opportunities of using this modality of exercise to promote physical activity among community-dwelling older adults. These findings can help inform program delivery and future research. The results offer some insight into the physical and psychosocial health benefits, and challenges, for insufficiently active community-dwelling older adults using kettlebells; enjoyable social interaction, the fun of learning something new and varied, the gratification from progress, improvement in the unwanted symptoms of knee arthritis and back pain, and enhanced self-perceptions of strength, appearance, and physical capacity, as well as the challenge of overcoming physical limitations, and training at high intensity. Sickness, medical procedures, and muscle soreness had negligible impact on attendance rates, and compliance with home training remained high, even though COVID-19 restrictions noticeably reduced motivation to train at home. Participants perceived group training to be a friendly, supportive, and encouraging environment, where they could meet like-minded people of a similar age and make new friends. For some participants, it was a respite from challenges with mental health. In the following sections we discuss the results in the context of existing literature, make recommendations for group-based community kettlebell programs for older adults, and consider directions for further investigation.

Many, but not all, of the findings in the present study, echo those of the ‘GOAL’ trial ^(25)^, which involved group exercise performed for 50-60 minutes three times weekly, for three to six months. The similarities in findings between the current study and the GOAL trial included: i) enjoyment and engagement were fostered by social connection, ii) post-exercise interaction (an embedded component of both trials) enriched social connections, iii) self-perceptions of physical health and capacity were enhanced by group exercise, iv) high- and low-performing outliers had a preference to train with others of a similar physical capacity, v) training with similar-age peers was preferential to regular group exercise with mixed-age adults, and vi) dismay once the intervention ended. What was dissimilar in the experience of the BELL trial participants, was that gender composition was not reported to be an issue, with some participants describing how they *preferred* training with those of the opposite sex. A loss of affiliation with or enjoyment of the group’s activities, was also not reported by the high- and low-performing outliers in the BELL trial.

The perceived benefits of feeling stronger and fitter, enhanced perceptions of physical appearance, and increased self-confidence, were consistent with previous studies which highlight the role of exercise in positively influencing how older adults see themselves and their perceived physical capacity ^(25, 46-48)^. The physical health benefits of sustained exercise are well documented ^(49, 50)^, with strength and endurance being strong indicators of health and predictive of mortality ^(51)^. Participants described doing more around the home and engaging in activities which they had previously found difficult or avoided altogether. This is consistent with Dionigi and Cannon ^(52)^, who identified perceived strength to be associated with improvements in performing day-to-day activities, having more energy, greater self-confidence and motivation, and a strong sense of achievement and self-esteem. Improvements in cardiovascular fitness and strength are important outcomes for older adults however, Killingback et al. ^(23)^ suggest that improvements in the affective domains relating to fun, enjoyment, camaraderie, friendship, and community, are essential, and promoting these outcomes from physical activity and exercise is key to maximising the uptake of older adults into new programs ^(53)^. Taken together, results from this study suggest that group-based kettlebell training improved older adults’ perceptions of health and wellbeing, increased incidental activity, and enhanced their functional capacity to perform activities of daily living.

Participants described their involvement as having given them a sense of purpose and enjoying the variety of exercises kettlebell training had offered. They liked the challenge of pushing themselves hard and accomplishing things which they thought were unattainable. Participants said the physical and psychological achievements were empowering, often disconfirming their maladaptive beliefs and behaviours relating to symptoms associated with chronic health conditions, such as arthritis and back pain. Previous investigations have indicated that health concerns and pain have constrained participants’ engagement with exercise ^(25)^, factors typically considered barriers to remaining active in later life ^(54)^. Participants in the BELL trial did not describe pain and symptoms associated with chronic conditions as barriers to engagement. On the contrary, one male in his 70s who was living with cancer, frequently outperformed his younger counterparts, and a female with an inflammatory arthropathy frequently recorded some of the highest daily training loads in the group, for both females and males. Extending the years spent functionally independent allows older adults to continue to pursue activities of enjoyment, improving quality of life, and reducing the risk of depression and loneliness ^(55)^.

Leveraging a social support network, such as those provided by group-exercise, can enable someone living with pain to participate in and be able to enjoy physical activity and exercise despite their symptoms ^(56)^. Healthcare providers have an important role in promoting and prescribing physical activity and exercise as part of routine care ^(57)^, and this influential role includes helping people live well with pain. Thornton et al. ^(58)^ recommend clinicians lead by example, as this provides credibility and empathy for the challenges facing patients. This sentiment was echoed by participants in the present study, who said they felt encouraged and reassured by seeing the instructor going through the same struggles. For people living with painful chronic health conditions, the influence of exercise interventions on pain is equivocal, with mostly small to moderate and inconsistent effects ^(56)^, suggesting that exercise alone may have only small positive effects on disability and limited effects on coping in those with low back pain ^(57)^. The experience of participants in this study, suggests a greater reduction in back pain than previous studies using exercise and resistance training programs for the treatment of chronic low back pain ^(59, 60)^ however, the mediators and moderators of this change are unclear.

Participants living with painful long-term conditions overcame the challenges of unpleasant symptoms, describing characteristics of grit and determination. An increased motivation and improved self-efficacy in managing their physical health was reported to have come, in part, from the direction and guidance provided by the instructor, and the support and encouragement of their peers. Exercise in all its forms is used by those in pain as a coping strategy, however, people living with pain will greatly benefit from a healthcare provider who is able to give person-specific support, help them make sense of their symptoms, provide coping strategies, and facilitate them being able to engage with regular physical activity and meaningful activities; to flexibility persist ^(61)^. Lack of motivation and self-efficacy are factors likely to undermine older adults with pain from being able to engage in meaningful activities. Those living with pain however, do seek opportunities to be active, and want a high-quality interaction with a healthcare provider to help and support them overcome their challenges ^(62)^. There is considerable value in a healthcare provider being able to provide accurate information and practical guidance as a trusted source of information and an early first point of contact ^(63)^. When advice and guidance is conflicting, those with back pain are more likely to withdraw from social activities, confused and anxious ^(64)^. Results of this study suggest that group-based kettlebell training, with an appropriately experienced and enthusiastic instructor, can help people living with pain be able to flexibility persist, and live well despite their symptoms.

Social connection increased the participants enjoyment and engagement in the trial. Consistent with previous observations ^(65)^, these results suggest that adherence to regular physical activity boosts self-reported improvements in overall health and well-being, and likely contributed to the large training volume participants said they were so proud of. The training was frequently reported to be “*hard*”, but many participants described having feelings of reward and achievement because of their effort. Some participants thrived on the effort, volume, and intensity, to such an extent that they consistently performed much more than was asked of them, and far exceeded the instructor’s expectations of capacity. Biedenweg et al. ^(66)^ reported that programmes offering *insufficient* vigorous exercise could be a barrier to adherence among older women. The findings of this study are a contrast to those which suggests that older adults are less likely to prefer vigorous exercise ^(67)^. In the present study, it was unclear how much of the participants’ enthusiasm was due to characteristics of the participant ^(68)^, the program design, characteristics of the instructor, the mode of exercise, or other factors.

Social connection and enjoyment was most evident among participants who reported a personal bond with one or more training partners; one of the tenets of self-categorisation theory ^(25)^. This was also evident by one third of the participants who completed the trial, still training together independently 18 months after the intervention ended. Bennett et al. ^(25)^ suggests that when older adults perceive their capability as markedly dissimilar to their peers, this can undermine their connecting to the group and enjoyment of the program, however, that was not apparent in this study. There were marked differences in baseline physical capacity of the participants in the BELL trial, but those differences were not reported to have adversely effected participants’ engagement with or enjoyment of the training. On the contrary, the high achievers, who were typically more competitive, said they enjoyed being able to help those who did not find things as easy, and liked the challenge of keeping ahead of or chasing those they saw as rivals. This mentality among males has been described by Bredland et al. ^(69)^ who reported participants secretly enjoying doing better than the person next to them, and those individuals frequently tending to push themselves too hard as a result. Participants who struggled most due to limited capacity, appreciated seeing their peers doing better than them, saying that it gave them something to aim for.

Being part of a community and having a social network was a strong theme. Consistent with previous studies ^(25, 67)^, some participants suggested that training with people their own age, albeit a cohort which spanned two decades, made them feel more comfortable. Some participants however, typically those who either wanted to push themselves very hard or just take it very easy, said they might have preferred to train with people of a similar capacity to themselves. In contrast to other studies which have reported mixed experiences based on gender composition ^(25)^, gender was not reported to have influenced participants’ experience in the BELL trial, positively or negatively.

Engagement remained very high with participants largely undeterred by the interruption of face-to-face training. Participants also remained motivated, performing a high volume of exercise five days a week for more than 3-months. Numerous motivators and barriers to engaging with and continuing in programs of resistance programs have previously been reported, which differ between target groups ^(67)^. Consistent with Simek et al. ^(70)^, participants said they had been less motivated training at home on their own, but that did not prevent any of the participants achieving a ‘personal best’ on the final day of the intervention. Group exercise is an important motivational factor for continued exercise adherence ^(66, 71, 72)^, which increases self-efficacy, improves feelings of well-being, decreases psychological stress ^(73)^, and provides a source of support, enjoyment, and belonging ^(22, 74, 75)^. Results from this study align with a growing body of evidence which emphasises the importance of social interaction in developing a positive attitude towards physical activity among older adults ^(76-78)^, which is particularly valuable during periods of forced isolation. Although classes were pre-planned and home-training quite limited, participants enjoyed the variety and autonomy of being able to change program variables: kettlebell weight, sets and reps, rest periods, and vary techniques to suit them. This aligns with other studies indicating that exercise variety positively affects engagement and enjoyment ^(79, 80)^. The BELL trial format was not an optimal fit for all participants, with changes likely necessary for kettlebell training to be used most effectively with different target groups of older adults.

Participants described feeling supported by a competent, enthusiastic, and encouraging instructor who was motivating, and inspired confidence. The importance of the instructor in group exercise has previously been described for a variety of older adult groups, including those with chronic conditions ^(26, 72, 81, 82)^. Instructors who nurture positive beliefs, improve participant self-efficacy, competence ^(83)^ and adherence ^(75)^. Killingback et al. ^(23)^ identified ‘instructor personality, professionalism, and a humanised approach’, as key factors in helping older people maintain adherence to group-exercise programs in the long-term, as these helped participants feel cared for, and established a sense of belonging. Gluchowski et al. ^(26)^ suggest that in the moments when older people doubt their abilities and capacity, it is at these critical moments of uncertainty and insecurity that instructors must instil a sense of confidence. Results of this study further add the importance of an encouraging instructor who listens, participates in the training, and guides older adults to challenge themselves.

Echoing the work of Tulle and Palmer ^(84)^, participants in the present study were motivated to take part as an act of agency, to make a contribution to social change, and to challenge aging stereotypes. In addition, the themes of accountability in this study, to the instructor, to the group, and to the trial, are consistent with those described by Biedenweg et al. ^(66)^. Furthermore, remarkably similar characteristics were observed among the men in the group as those described by Bredland et al. ^(69)^; being less likely to engage in healthy aging activities to improve their balance, enjoying the opportunity to ‘flex their muscles’, hiding things they felt might be perceived as a physical weakness, and improvements from training making them feel more “*useful*” at home. Results of this study are also consistent with the work of Ericson et al. ^(85)^, who describe seven health resources important for the maintenance of health, and reasons for older women continuing to engage in resistance training: social relations and care, positive energy, self-worth, capability in and about physical activity, the habit of exercising, identity as an exercising person and womanhood.

The following statements and practical recommendations are based on observation of the intervention, and findings of the present study. Hardstyle kettlebell training can be used with inexperienced older adults in a group exercise setting. Kettlebell training is accessible to otherwise healthy older adults, regardless of previous training history, with all exercises modifiable. An optimal number of participants in a group setting is likely to be no more than ten, with the most desirable outcomes more likely achievable when participants have a similar physical capacity. Having multiple kettlebell weights, from 4-6 kg and increasing in increments of 2 kg, offers maximum flexibility in programming variables and participant’s choice of exercise, and it should be anticipated that some older adults will be able to deadlift bodyweight i.e., heavy kettlebells may be beneficial. Lack of resistance training history is likely to influence muscle soreness, its severity, and impact, with progression of loading best managed accordingly. Older adults’ physical capacity and/or desire may initially exceed their comfortable tolerance for physiological adaptation, with “easy” low-load, high-repetition exercises still likely to cause muscle soreness. Structure, exercise variety, clear goal setting, and progress tracking are strong motivators. Technique acquisition may take longer for older adults, but this may not need hinder or delay progress. Ideal mobility may not be achievable, but this also may not need to hinder or delay progress. Hardstyle training provides instructors with a practice and programming framework. Knowledge of hardstyle practices, competency in performing the exercises, and experience in delivery, are desirable, with the ‘instructor’ likely to be an influential program variable in participant engagement and attainment of outcomes.

Despite the strengths of this study, it is not without limitations. First, while a well-established relationship between the examiner and participants throughout the intervention and the present study facilitated open and frank discussion, bias may have positively skewed participants’ responses. Second, the researcher’s role in knowledge production is at the heart of reflexive thematic analysis. Themes evoke participants voices, but ultimately tell the researcher’s story about the data, which is a subjective interpretation developed with theoretical assumptions, which have been made through the lens of a particular social, cultural, disciplinary, and ideological position. Finally, participants represented only one ethnic group: older Caucasian Australians. The likelihood however that these results may be transferable to other insufficiently active community-living older adults, is enhanced by thematic saturation and the heterogenous representation of physical capacity, exercise experience, and health conditions within the cohort.

Several steps were taken to maximise the research quality ^(86)^. Firstly, the purposive sample; including all participants who commenced the training, including those who dropped out or were unable to fully complete the intervention period, allowed us to present a meaningful representation of the participants’ experiences of the training program, both positive and negative. Secondly, ongoing communication with the participants who continued to train together after the trial, allowed the lead author to be fully immersed in and report upon the lived experience of participants which provides rigor. Thirdly, research sincerity is enhanced by transparency of methods and self-reflexibility around researcher roles and biases. Finally, we provide a discussion which interconnects the literature providing a meaningful coherence of results.

Future studies of hardstyle kettlebell training with older adults should investigate a) the experience of smaller group trials conducted within a clinic or community-setting, b) effectiveness of a similar intervention provided to participants with a more homogenous physical capacity, c) longer intervention periods with lower intensity and lower volume, d) single case experimental designs for common musculoskeletal conditions, e) suitability for older adults not living independently, f) effects on knee arthritis, g) effects on persistent non-specific low back pain, g) effects for mental health disorders, and h) effects for reducing falls risk.

In summary, the experiences of participants in the BELL trial, reveal the efficacy and perceived health benefits associated with group-based hardstyle kettlebell training with peers of a similar age, and highlight some of the challenges of delivering a group-based kettlebell program that meets the diverse needs of older men and women. Group training provided a valued opportunity for participants to build a new social support network, which greatly contributed to the participant’s enjoyment of the training and high rate of engagement, with the instructor likely to be influential in optimising outcomes. Age, training status, sex, and health condition did not negatively impact engagement or participation in the training. Training performance for participants living with a chronic health condition or persistent pain, challenged their self-schema, often disconfirming maladaptive beliefs and behaviours. Participants’ experiences in this large-scale replication of a successful clinic-based group-exercise program, indicate that a pragmatic approach to hardstyle kettlebell training may be an effective means of engaging insufficiently active older adults in regular resistance-based exercise within the community, with minimal barriers to entry. These results corroborated earlier studies highlighting the value of the social features of group-exercise, in fostering a supportive social environment to optimise participants’ health and wellbeing. Further research is warranted to optimise program design and delivery within community settings, to test subgroups of older adults in a clinical framework, and to identify strategies which support long-term engagement.

## Supporting information

COREQ checklist

SurveyMonkey questionnaire - your kettlebell experience

SurveyMonkey questionnaire - positive and beneficial effects

SurveyMonkey - negative and undesirable effects

## Data Availability

Data will be available in accordance with Bond University Research Data Management and Sharing Policy (TLR 5.12)

## Author note

Neil Meigh is a doctoral candidate at Bond University’s Institute of Health and Sport holding a Doctor of Physiotherapy and Bachelor of Exercise Science. He is a former Physiotherapy clinic owner where he ran small group kettlebell classes using the training practices which were used in the BELL trial, which he conducted.

Alexandra Davidson is a doctoral candidate at Bond University’s Faculty of Health Sciences and Medicine. She has a Master of Nutrition & Dietetic Practice and Bachelor of Biomedical Science. As an Accredited Practicing Dietitian, her qualitative research is investigating interprofessional collaboration in general practice.

Dr Justin Keogh is an Associate Professor of Sport and Exercise Science at Bond University. His research interests are as an Exercise Scientist investigating the benefits of exercise, particularly resistance training in cancer survivors and older adults, and improving athletic performance. Dr Keogh is a Fellow of the Australian Association of Gerontology and serves on the Australian and New Zealand Society of Sarcopenia and Frailty Research’s Sarcopenia Diagnosis Task Force Committee.

Dr Wayne Hing is a Professor of Physiotherapy at Bond University who has supervised over 70 doctoral students to completion.

## Supplementary data

This project will contain the following supplementary data:

- COREQ Checklist_BELL.docx
- Survey Monkey Questionnaire_your kettlebell experience.docx
- Survey Monkey Questionnaire_positive and beneficial effects.docx
- Survey Monkey Questionnaire_negative and undesirable effects.docx

Extended data are available under the terms of the Creative Commons Zero “No rights reserved” data waiver (CC0 1.0 Public domain dedication).

## References

1. Statistics ABo. Life expectancy continues to increase in Australia: Australian Government; 2020 [Available from: http://ow.ly/drTE50EERF2.

2. Welfare AIoHa. Health of older people [Report]. Australian Government; 2020 [updated 23 July 2020. Available from: https://www.aihw.gov.au/reports/australias-health/health-of-older-people.

3. Salomon JA, Wang H, Freeman MK, Vos T, Flaxman AD, Lopez AD, et al. Healthy life expectancy for 187 countries, 1990–2010: a systematic analysis for the Global Burden Disease Study 2010. The Lancet. 2012;380(9859):2144–62.

4. Paterson DH, Warburton DER. Physical activity and functional limitations in older adults: a systematic review related to Canada’s Physical Activity Guidelines. International Journal of Behavioral Nutrition and Physical Activity. 2010;7(1):38.

5. Daskalopoulou C, Stubbs B, Kralj C, Koukounari A, Prince M, Prina AM. Physical activity and healthy ageing: a systematic review and meta-analysis of longitudinal cohort studies. Ageing research reviews. 2017;38:6–17.

6. Ashdown-Franks G, Firth J, Carney R, Carvalho AF, Hallgren M, Koyanagi A, et al. Exercise as medicine for mental and substance use disorders: a meta-review of the benefits for neuropsychiatric and cognitive outcomes. Sports Med. 2020;50(1):151–70.

7. Levin O, Netz Y, Ziv G. The beneficial effects of different types of exercise interventions on motor and cognitive functions in older age: a systematic review. Eur Rev Aging Phys Act. 2017;14(1):20.

8. Kaushal N, Desjardins-Crépeau L, Langlois F, Bherer L. The effects of multi-component exercise training on cognitive functioning and health-related quality of life in older adults. Int J Behav Med. 2018;25(6):617–25.

9. Raafs BM, Karssemeijer EGA, Horst Lvd, Aaronson JA, Olde Rikkert MGM, Kessels RPC. Physical exercise training improves quality of life in healthy older adults: a meta-analysis. J Aging Phys Act. 2020;28(1):81–93.

10. Netz Y. Is there a preferred mode of exercise for cognition enhancement in older age? - a narrative review. Frontiers in Medicine. 2019;6(57).

11. Tse AC, Wong TW, Lee PH. Effect of low-intensity exercise on physical and cognitive health in older adults: a systematic review. Sports Med Open. 2015;1(1):37.

12. Peterson MD, Rhea MR, Sen A, Gordon PM. Resistance exercise for muscular strength in older adults: a meta-analysis. Ageing research reviews. 2010;9(3):226–37.

13. Lewis FJ, Stewart HC, Roddam H. Effects of exercise interventions on physical function, mobility, frailty status and strength in the prefrail population: a review of the evidence base for practice. European journal of physiotherapy. 2019:1–9.

14. Chen F-T, Hopman RJ, Huang C-J, Chu C-H, Hillman CH, Hung T-M, et al. The effect of exercise training on brain structure and function in older adults: a systematic review based on evidence from randomized control trials. Journal of clinical medicine. 2020;9(4):914.

15. Samitz G, Egger M, Zwahlen M. Domains of physical activity and all-cause mortality: systematic review and dose–response meta-analysis of cohort studies. Int J Epidemiol. 2011;40(5):1382–400.

16. Borde R, Hortobagyi T, Granacher U. Dose-response relationships of resistance training in healthy old adults: a systematic review and meta-analysis. Sports Med. 2015;45(12):1693–720.

17. Hallal PC, Andersen LB, Bull FC, Guthold R, Haskell W, Ekelund U. Global physical activity levels: surveillance progress, pitfalls, and prospects. Lancet. 2012;380(9838):247–57.

18. Lazarus N, Izquierdo M, Higginson I, D.R. Harridge S. Exercise deficiency diseases of ageing: The primacy of exercise and muscle strengthening as first line therapeutic agents to combat frailty 2018.

19. Compernolle S, De Cocker K, Cardon G, De Bourdeaudhuij I, Van Dyck D. Older adults’ perceptions of sedentary behavior: a systematic review and thematic synthesis of qualitative studies. The Gerontologist. 2019;60(8):e572–e82.

20. Chase J-AD. Interventions to increase physical activity among older adults: a meta-analysis. The Gerontologist. 2015;55(4):706–18.

21. Hong S-Y, Hughes S, Prohaska T. Factors affecting exercise attendance and completion in sedentary older adults: a meta-analytic approach. Journal of physical activity and health. 2008;5(3):385–97.

22. Farrance C, Tsofliou F, Clark C. Adherence to community based group exercise interventions for older people: a mixed-methods systematic review. Prev Med. 2016;87:155–66.

23. Killingback C, Tsofliou F, Clark C. Older people’s adherence to community-based group exercise programmes: a multiple-case study. BMC Public Health. 2017;17(1):115-.

24. Reichstadt J, Sengupta G, Depp CA, Palinkas LA, Jeste DV. Older adults’ perspectives on successful aging: qualitative interviews. The American journal of geriatric psychiatry. 2010;18(7):567–75.

25. Bennett EV, Hurd Clarke L, Wolf SA, Dunlop WL, Harden SM, Liu Y, et al. Older adults’ experiences of group-based physical activity: A qualitative study from the ‘GOAL’ randomized controlled trial. Psychol Sport Exerc. 2018;39:184–92.

26. Gluchowski A, Warbrick I, Oldham T, Harris N. ‘I have a renewed enthusiasm for going to the gym’: what keeps resistance-trained older adults coming back to the gym? Qualitative Research in Sport, Exercise and Health. 2018:1–13.

27. Marcelino C. Effects of kettlebell training on functional performance, postural stability and isokinetic strength of lower limbs in individuals with Parkinson’s disease [masters]. Brazil: Univeristy of Brazil. 2017.

28. Chen HT, Wu HJ, Chen YJ, Ho SY, Chung YC. Effects of 8-week kettlebell training on body composition, muscle strength, pulmonary function, and chronic low-grade inflammation in elderly women with sarcopenia. Exp Gerontol. 2018;112:112–8.

29. Jay K, Jakobsen MD, Sundstrup E, Skotte JH, Jorgensen MB, Andersen CH, et al. Effects of kettlebell training on postural coordination and jump performance: a randomized controlled trial. J Strength Cond Res. 2013;27(5):1202–9.

30. Meigh NJ, Keogh JWL, Schram B, Hing W, Rathbone EN. Effects of supervised high-intensity hardstyle kettlebell training on grip strength and health-related physical fitness in insufficiently active older adults: The BELL pragmatic controlled trial. medRxiv. 2021:2021.06.27.21259191.

31. Tong A, Sainsbury P, Craig J. Consolidated criteria for reporting qualitative research (COREQ): a 32-item checklist for interviews and focus groups. Int J Qual Health Care. 2007;19(6):349–57.

32. Robinson RS. Purposive Sampling. In: Michalos AC, editor. Encyclopedia of Quality of Life and Well-Being Research. Dordrecht: Springer Netherlands; 2014. p. 5243–5.

33. Trint. AI audio transcription software. 2020.

34. QSR_International. NVivo qualitative data analysis software. Version 12 ed2018.

35. Lingard L. Beyond the default colon: effective use of quotes in qualitative research. Perspectives on medical education. 2019;8(6):360–4.

36. Kvale S, Brinkmann S. Interviews: Learning the craft of qualitative research interviewing: SAGE; 2009.

37. Archer M, Bhaskar R, Collier A, Lawson T, Norrie A. Critical realism: essential readings: Routledge; 2013.

38. Fryer T. Conceptualising graduate outcomes with critical realism. Higher Education Policy. 2021:1–16.

39. Braun V, Clarke V, Weate P. Using thematic analysis in sport and exercise research. Routledge handbook of qualitative research in sport and exercise. 2016:191–205.

40. Braun V, Clarke V. Reflecting on reflexive thematic analysis. Qualitative Research in Sport, Exercise and Health. 2019;11(4):589–97.

41. Braun V, Clarke V. One size fits all? What counts as quality practice in (reflexive) thematic analysis? Qualitative research in psychology. 2020:1–25.

42. Braun V, Clarke V. Can I use TA? Should I use TA? Should I not use TA? Comparing reflexive thematic analysis and other pattern-based qualitative analytic approaches. Counselling and Psychotherapy Research. 2021;21(1):37–47.

43. Fletcher AJ. Applying critical realism in qualitative research: methodology meets method. International Journal of Social Research Methodology. 2017;20(2):181–94.

44. Adu P. Qualitative analysis coding and categorizing Online 2013 [Available from: https://www.slideshare.net/kontorphilip/qualitative-analysis-coding-and-categorizing.

45. Phoenix C, Orr N. Analysing exceptions within qualitative data: promoting analytical diversity to advance knowledge of ageing and physical activity. Qualitative Research in Sport, Exercise and Health. 2017;9(3):271–84.

46. Bennett EV, Clarke LH, Kowalski KC, Crocker PR. From pleasure and pride to the fear of decline: exploring the emotions in older women’s physical activity narratives. Psychol Sport Exerc. 2017;33:113–22.

47. Liechty T, Ribeiro NF, Sveinson K, Dahlstrom L. “It’s about what I can do with my body”: body image and embodied experiences of aging among older Canadian men. International Journal of Men’s Health. 2014;13(1).

48. von Berens Å, Koochek A, Nydahl M, Fielding R, Gustafsson T, Kirn D, et al. “Feeling more self-confident, cheerful and safe”. experiences from a health-promoting intervention in community dwelling older adults - a qualitative study. J Nutr Health Aging. 2018;22(4):541–8.

49. Chodzko-Zajko WJ, Proctor DN, Singh MAF, Minson CT, Nigg CR, Salem GJ, et al. Exercise and physical activity for older adults. Med Sci Sports Exerc. 2009;41(7):1510–30.

50. Bangsbo J, Blackwell J, Boraxbekk C-J, Caserotti P, Dela F, Evans AB, et al. Copenhagen Consensus statement 2019: physical activity and ageing. Br J Sports Med. 2019;53(14):856–8.

51. Strassmann A, Steurer-Stey C, Lana KD, Zoller M, Turk AJ, Suter P, et al. Population-based reference values for the 1-min sit-to-stand test. International Journal of Public Health. 2013;58(6):949–53.

52. Dionigi RA, Cannon J. Older adults’ perceived changes in physical self-worth associated with resistance training. Res Q Exerc Sport. 2009;80(2):269–80.

53. Brawley LR, Latimer AE. Physical activity guides for Canadians: messaging strategies, realistic expectations for change, and evaluation. Appl Physiol Nutr Metab. 2007;32(S2E):S170–S84.

54. Franco MR, Tong A, Howard K, Sherrington C, Ferreira PH, Pinto RZ, et al. Older people’s perspectives on participation in physical activity: a systematic review and thematic synthesis of qualitative literature. Br J Sports Med. 2015;49(19):1268–76.

55. Aartsen M, Jylhä M. Onset of loneliness in older adults: results of a 28 year prospective study. European journal of ageing. 2011;8(1):31–8.

56. Vader K, Patel R, Doulas T, Miller J. Promoting participation in physical activity and exercise among people living with chronic pain: a qualitative study of strategies used by people with pain and their recommendations for health care providers. Pain Med. 2020;21(3):625–35.

57. Orchard J. Prescribing and dosing exercise in primary care. Australian Journal for General Practitioners. 2020;49:182–6.

58. Thornton JS, Frémont P, Khan K, Poirier P, Fowles J, Wells GD, et al. Physical activity prescription: a critical opportunity to address a modifiable risk factor for the prevention and management of chronic disease: a position statement by the Canadian Academy of Sport and Exercise Medicine. Br J Sports Med. 2016;50(18):1109–14.

59. Searle A, Spink M, Ho A, Chuter V. Exercise interventions for the treatment of chronic low back pain: a systematic review and meta-analysis of randomised controlled trials. Clin Rehabil. 2015;29(12):1155–67.

60. Tataryn N, Simas V, Catterall T, Furness J, Keogh JWL. Posterior-Chain Resistance Training Compared to General Exercise and Walking Programmes for the Treatment of Chronic Low Back Pain in the General Population: A Systematic Review and Meta-Analysis. Sports Medicine - Open. 2021;7(1):17.

61. Lennox Thompson B, Gage J, Kirk R. Living well with chronic pain: a classical grounded theory. Disabil Rehabil. 2020;42(8):1141–52.

62. Karlsson L, Gerdle B, Takala EP, Andersson G, Larsson B. Experiences and attitudes about physical activity and exercise in patients with chronic pain: a qualitative interview study. J Pain Res. 2018;11:133–44.

63. Cridland K, Pritchard S, Rathi S, Malliaras P. ‘He explains it in a way that I have confidence he knows what he is doing’: A qualitative study of patients’ experiences and perspectives of rotator-cuff-related shoulder pain education. Musculoskeletal Care.n/a(n/a).

64. Alhowimel A, Alotaibi M, Coulson N, Radford K. Psychosocial consequences of diagnosing nonspecific low-back pain radiologically: a qualitative study. Physiother Theory Pract. 2020:1–7.

65. Holt-Lunstad J, Smith TB, Layton JB. Social relationships and mortality risk: a meta-analytic review. PLoS Med. 2010;7(7):e1000316.

66. Biedenweg K, Meischke H, Bohl A, Hammerback K, Williams B, Poe P, et al. Understanding older adults’ motivators and barriers to participating in organized programs supporting exercise behaviors. The journal of primary prevention. 2014;35(1):1–11.

67. Burton NW, Khan A, Brown WJ. How, where and with whom? Physical activity context preferences of three adult groups at risk of inactivity. Br J Sports Med. 2012;46(16):1125–31.

68. Pocnet C, Popp J, Jopp D. The power of personality in successful ageing: a comprehensive review of larger quantitative studies. European journal of ageing. 2020.

69. Bredland EL, Söderström S, Vik K. Challenges and motivators to physical activity faced by retired men when ageing: a qualitative study. BMC Public Health. 2018;18(1):627–9.

70. Simek EM, McPhate L, Haines TP. Adherence to and efficacy of home exercise programs to prevent falls: a systematic review and meta-analysis of the impact of exercise program characteristics. Prev Med. 2012;55(4):262–75.

71. Appleby KM, Dieffenbach K. “Older and faster”: exploring elite masters cyclists’ involvement in competitive sport. The Sport Psychologist. 2016;30(1):13–23.

72. Fisken AL, Waters DL, Hing WA, Keogh JW. Perceptions towards aqua-based exercise among older adults with osteoarthritis who have discontinued participation in this exercise mode. Australas J Ageing. 2016;35(1):12–7.

73. McAuley E, Blissmer B, Katula J, Duncan TE. Exercise environment, self-efficacy, and affective responses to acute exercise in older adults. Psychology and Health. 2000;15(3):341–55.

74. Hartley SE, Yeowell G. Older adults’ perceptions of adherence to community physical activity groups. Ageing and Society. 2015;35(8):1635–56.

75. Hawley-Hague H, Horne M, Skelton DA, Todd C. Older adults’ uptake and adherence to exercise classes: instructors’ perspectives. J Aging Phys Act. 2016;24(1):119–28.

76. Bidonde MJ, Goodwin DL, Drinkwater DT. Older women’s experiences of a fitness program: the importance of social networks. J Appl Sport Psychol. 2009;21(1):S86–S101.

77. Grant BC. An insider’s view on physical activity in later life. Psychol Sport Exerc. 2008;9(6):817–29.

78. Lozano-Sufrategui L, Pringle A, Carless D, McKenna J. ‘It brings the lads together’: a critical exploration of older men’s experiences of a weight management programme delivered through a Healthy Stadia project. Sport Soc. 2017;20(2):303–15.

79. Sylvester BD, Lubans DR, Eather N, Standage M, Wolf SA, McEwan D, et al. Effects of variety support on exercise-related well-being. Applied Psychology: Health and Well-Being. 2016;8(2):213–31.

80. Sylvester BD, Standage M, McEwan D, Wolf SA, Lubans DR, Eather N, et al. Variety support and exercise adherence behavior: experimental and mediating effects. J Behav Med. 2016;39(2):214–24.

81. Bundon A, Clarke LH, Miller WC. Frail older adults and patterns of exercise engagement: understanding exercise behaviours as a means of maintaining continuity of self. Qualitative research in sport, exercise and health. 2011;3(1):33–47.

82. Janssen SL, Stube JE. Older adults’ perceptions of physical activity: a qualitative study. Occup Ther Int. 2014;21(2):53–62.

83. Cross SE, Markus HR. Self-schemas, possible selves, and competent performance. J Educ Psychol. 1994;86(3):423.

84. Tulle E, Palmer C. Engaging participants in qualitative research: methodological reflections on studying active older lives in Scotland and Australia. Qualitative Research in Sport, Exercise and Health. 2020:1–15.

85. Ericson H, Quennerstedt M, Skoog T, Johansson M. Health resources, ageing and physical activity: a study of physically active women aged 69-75 years. Qualitative Research in Sport, Exercise and Health. 2018;10(2):206–22.

86. Tracy SJ. Qualitative quality: Eight “big-tent” criteria for excellent qualitative research. Qualitative inquiry. 2010;16(10):837–51.

