## Supplementary material for "“If somebody had told me I’d feel like I do now, I wouldn’t have believed them…” Older adults’ experiences of the BELL trial: a qualitative study": COREQ checklist

### The BELL Trial: Consolidated criteria for reporting qualitative studies (COREQ): 32-item checklist.

| No. | Item | Guide questions / description | Page |
|---|---|---|---|
| **Domain 1: Research team and reflexivity** | | | |
| *Personal Characteristics* | | | |
| 1. | Interviewer/facilitator | Which author/s conducted the interview or focus group? | 2 |
| 2. | Credentials | What were the researcher's credentials? e.g., PhD, MD | 8 |
| 3. | Occupation | What was their occupation at the time of the study? | 8 |
| 4. | Gender | Was the researcher male or female? | 2 |
| 5. | Experience and training | What experience or training did the researcher have? | 8 |
| *Relationship with participants* | | | |
| 6. | Relationship established | Was a relationship established prior to study commencement? | 2, 8 |
| 7. | Participant knowledge of the interviewer | What did the participants know about the researcher? *e.g., personal goals, reasons for doing the research* | 2, 8 |
| 8. | Interviewer characteristics | What characteristics were reported about the interviewer/facilitator? *e.g., bias, assumptions, reasons and interests in the research topic* | 8 |
| **Domain 2: study design** | | | |
| *Theoretic framework* | | | |
| 9. | Methodological orientation and Theory | What methodological orientation was stated to underpin the study? *e.g., grounded theory, discourse analysis, ethnography, phenomenology, content analysis* | 3 |
| *Participant selection* | | | |
| 10. | Sampling | How were participants selected? *e.g., purposive, convenience,* consecutive, snowball | 2 |
| 11. | Method of approach | How were participants approached? *e.g., face-to-face, telephone, mail, email* | 2 |
| 12. | Sample size | How many participants were in the study? | 2 |
| 13. | Non-participation Setting | How many people refused to participate or dropped out? Reasons? | 2 |
| *Setting* | | | |
| 14. | Setting of data collection | here was the data collected? *e.g., home, clinic, workplace* | 2 |
| 15. | Presence of non-participants | Was anyone else present besides the participants and researchers? | 2 |
| 16. | Description of sample | What are the important characteristics of the sample*? e.g., demographic data, date* | 2 |
| *Data collection* | | | |
| 17. | Interview guide | Were questions, prompts, guides provided by the authors? Was it pilot tested? | 2 |
| 18. | Repeat interviews | Were repeat interviews carried out? If yes, how many? | 2 |
| 19. | Audio/visual recording | Did the research use audio or visual recording to collect the data? | 2 |
| 20. | Field notes | Were field notes made during and/or after the interview or focus group? | 2 |
| 21. | Duration | What was the duration of the interviews or focus group? | 2 |
| 22. | Data saturation | Was data saturation discussed? | 8 |
| 23. | Transcripts returned | Were transcripts returned to participants for comment and/or correction? | 2 |
| **Domain 3: analysis and findings** | | | |
| *Data analysis* | | | |
| 24. | Number of data coders | How many data coders coded the data? | 3 |

| | | | |
|---|---|---|---|
| 25. | Description of the coding tree | Did authors provide a description of the coding tree? | n/a |
| 26. | Derivation of themes | Were themes identified in advance or derived from the data? | 3 |
| 27. | Software | What software, if applicable, was used to manage the data? | 2 |
| 28. | Participant checking | Did participants provide feedback on the findings? | 2 |
| *Reporting* | | | |
| 29. | Quotations presented | Were participant quotations presented to illustrate the themes / findings? Was each quotation identified? *e.g., participant number* | 3-6 |
| 30. | Data and findings consistent | Was there consistency between the data presented and the findings? | 3-6 |
| 31. | Clarity of major themes | Were major themes clearly presented in the findings? | 3-6 |
| 32. | Clarity of minor themes | Is there a description of diverse cases or discussion of minor themes? | 6-8 |
