## Supplementary material for "“If somebody had told me I’d feel like I do now, I wouldn’t have believed them…” Older adults’ experiences of the BELL trial: a qualitative study": SurveyMonkey questionnaire - your kettlebell experience

### Older adults’ experiences of supervised hardstyle kettlebell training: A qualitative study from the BELL pragmatic controlled trial

**SurveyMonkey questionnaire:** *Your kettlebell experience*

| **How strongly do you agree that the following were motivators for wanting to participate in the trial?** | Strongly agree | Disagree | Neither agree nor disagree | Agree | Strongly agree |
|---|---|---|---|---|---|
| To lose weight | ○ | ○ | ○ | ○ | ○ |
| To increase physical activity | ○ | ○ | ○ | ○ | ○ |
| To try something new | ○ | ○ | ○ | ○ | ○ |
| To keep independent and healthy | ○ | ○ | ○ | ○ | ○ |
| Having a reason to leave your home | ○ | ○ | ○ | ○ | ○ |
| To meet new people | ○ | ○ | ○ | ○ | ○ |
| To have a weekly routine | ○ | ○ | ○ | ○ | ○ |
| Recommended by friend or family member | ○ | ○ | ○ | ○ | ○ |
| To support someone else | ○ | ○ | ○ | ○ | ○ |
| To challenge yourself | ○ | ○ | ○ | ○ | ○ |
| You were previously very physically active | ○ | ○ | ○ | ○ | ○ |
| To prevent a chronic health condition e.g., heart disease | ○ | ○ | ○ | ○ | ○ |
| To be involved in a university research project | ○ | ○ | ○ | ○ | ○ |
| To 'give back' and help the community | ○ | ○ | ○ | ○ | ○ |
| Wanting to learn more about healthy ageing | ○ | ○ | ○ | ○ | ○ |

| **How strongly do you agree with the following statements?** | Strongly agree | Disagree | Neither agree nor disagree | Agree | Strongly agree |
|---|---|---|---|---|---|
| I enjoyed training with kettlebells | ○ | ○ | ○ | ○ | ○ |
| Kettlebell training has improved my physical health and/or capacity | ○ | ○ | ○ | ○ | ○ |
| Kettlebell training has improved my mental health and/or sense of wellbeing | ○ | ○ | ○ | ○ | ○ |
| I feel confident that I can train at home on my own with a kettlebell | ○ | ○ | ○ | ○ | ○ |
| I will continue using a kettlebell for exercise at home after the trial | ○ | ○ | ○ | ○ | ○ |
| What I've physically able to achieve exceeded my expectations | ○ | ○ | ○ | ○ | ○ |
| Regular physical activity or sport has played an important role in my life | ○ | ○ | ○ | ○ | ○ |
| I enjoyed exercising at home | ○ | ○ | ○ | ○ | ○ |
| The extra home exercises were helpful | ○ | ○ | ○ | ○ | ○ |

| **How important were the following to your overall enjoyment of the training** | Unimportant | Somewhat important | Moderately important | Very important | Extremely important |
|---|---|---|---|---|---|
| Variety of exercises | ○ | ○ | ○ | ○ | ○ |
| The instructor's personality | ○ | ○ | ○ | ○ | ○ |
| Social interaction and connecting with others | ○ | ○ | ○ | ○ | ○ |
| Exercising as a group | ○ | ○ | ○ | ○ | ○ |
| Having a weekly routine and structure | ○ | ○ | ○ | ○ | ○ |
| The format of the training sessions | ○ | ○ | ○ | ○ | ○ |
| Having set goals and reaching targets | ○ | ○ | ○ | ○ | ○ |
| Other (please specify): | | | | | |

| **How aware have you been of the following** | Not at all aware | Slightly aware | Moderately aware | Very aware | Extremely aware |
|---|---|---|---|---|---|
| Having a stronger and more enduring grip | ○ | ○ | ○ | ○ | ○ |
| Feeling less 'puffed' when walking quickly or ascending stairs | ○ | ○ | ○ | ○ | ○ |
| Having stronger legs | ○ | ○ | ○ | ○ | ○ |
| Being able to rise from a chair or seat quicker with less difficulty | ○ | ○ | ○ | ○ | ○ |
| Feeling less generalised physical fatigue | ○ | ○ | ○ | ○ | ○ |
| Physical changes i.e., less body fat or more muscle tone | ○ | ○ | ○ | ○ | ○ |
| Getting down to the ground and back up again with less difficulty | ○ | ○ | ○ | ○ | ○ |
| Lifting heavier objects around the home | ○ | ○ | ○ | ○ | ○ |
| Feeling less concerned about lifting or manipulating heavy objects | ○ | ○ | ○ | ○ | ○ |
| Feeling steadier and more balanced on your feet | ○ | ○ | ○ | ○ | ○ |
| Feeling more disciplined having a regular routine of exercise | ○ | ○ | ○ | ○ | ○ |
| Improvements in a chronic health condition such as back or knee conditions | ○ | ○ | ○ | ○ | ○ |
| Weight loss | ○ | ○ | ○ | ○ | ○ |
| Improved sleep | ○ | ○ | ○ | ○ | ○ |
| More energy | ○ | ○ | ○ | ○ | ○ |
| Feeling more cheerful or happy | ○ | ○ | ○ | ○ | ○ |
| Feeling more positive or encouraged | ○ | ○ | ○ | ○ | ○ |
| Improved wellbeing | ○ | ○ | ○ | ○ | ○ |
| Feeling a sense of achievement | ○ | ○ | ○ | ○ | ○ |

**Has your physical performance in the trial exceeded your prior expectations?**

**When you trained at home, did you train alone or was someone there to help or train with you?**

**During the trial, did you show or teach someone else how to use a kettlebell?**

**Have family or friends expressed interest in your participation in the trial and/or you training with a kettlebell?** (If 'Yes', please explain:)

**If the kettle bell training could have been designed and delivered just for you, what would it have looked like i.e., how would it have been different?**
