## Supplementary material for "“If somebody had told me I’d feel like I do now, I wouldn’t have believed them…” Older adults’ experiences of the BELL trial: a qualitative study": SurveyMonkey questionnaire - positive and beneficial effects

### Older adults’ experiences of supervised hardstyle kettlebell training: A qualitative study from the BELL pragmatic controlled trial

**SurveyMonkey questionnaire:** *Positive and beneficial effects*

1. Describe any physical affects you have experienced from kettlebell training which you feel have been positive or beneficial.
2. Describe any psycho-social affects you have experienced from kettlebell training which you feel have been positive or beneficial.
3. What 3 things have you enjoyed the most?
   i.
   ii.
   iii.
4. What would you say are the benefits of kettlebell training?
5. What did you find to be especially easy, achievable, and motivating? (multiple responses allowed)
   a. Easy
   b. Achievable
   c. Motivating

| | Definitely not | Probably not | Unsure | Probably yes | Definitely yes |
|---|---|---|---|---|---|
| 6. Would you recommend kettlebell training to your friends of a similar age? | ○ | ○ | ○ | ○ | ○ |

| | Very unlikely | Unlikely | Neutral | Likely | Very likely |
|---|---|---|---|---|---|
| 7. If similar training was available in the community, how likely is it that you would attend? | ○ | ○ | ○ | ○ | ○ |
