## Supplementary material for "“If somebody had told me I’d feel like I do now, I wouldn’t have believed them…” Older adults’ experiences of the BELL trial: a qualitative study": SurveyMonkey - negative and undesirable effects

### Older adults’ experiences of supervised hardstyle kettlebell training: A qualitative study from the BELL pragmatic controlled trial

**SurveyMonkey questionnaire:** *Negative and undesirable effects*

1. Describe any physical affects you have experienced from kettlebell training which you feel have been negative (specifically including injury, but excluding DOMS) or undesirable/harmful
2. Describe any psycho-social affects you have experienced from kettlebell training which you feel have been negative or undesirable/harmful
3. Are there any reasons that you would not recommend kettlebell training to other people of your age and background?
4. What 3 things have you least enjoyed about kettlebell training; either being a participant in the group, or specifically related to training with kettlebells?
   - i.
   - ii.
   - iii.
5. What would you say are the potential harms of kettlebell training?
6. What did you find to be especially hard, unachievable, and demotivating? (multiple responses allowed).
   a. Hard
   b. Unachievable
   c. Demotivating
